# Dissecting the genetic architecture of knee alignment reveals its contribution to osteoarthritis risk

**DOI:** 10.64898/2026.06.23.26356332

**Authors:** Mijin Jung, Faten Alomar, Clarissa R. Coveney, Shibo Chen, Sarah E Orr, Jolet Mimpen, Marina Nikolic, Kaitlyn A Flynn, Yuandan Zhang, Raja Ebsim, Fiona R Saunders, Jennifer S Gregory, Richard M Aspden, Nicholas C Harvey, Claudia Lindner, Simon GF Abram, Chrissy Hammond, George Davey Smith, Eleftheria Zeggini, Sarah Snelling, Terence D. Capellini, Sarah J Rice, John P. Kemp, Jonathan H Tobias, Tim Cootes, Benjamin G Faber

## Abstract

**Objectives:** To investigate the biological and clinical relevance of knee alignment in osteoarthritis by integrating population-scale imaging, genome-wide association, and functional genetic analyses.

**Methods:** Femorotibial angle was derived from dual-energy X-ray absorptiometry scans in UK Biobank using machine-learning methods. Associations with knee and hip osteoarthritis outcomes were assessed. A genome-wide association study of mean femorotibial angle was performed, followed by fine-mapping and pathway enrichment analyses. Mendelian randomization was used to explore potential causal relationships.

**Results:** Varus alignment was strongly and progressively associated with knee pain, knee osteoarthritis, and total knee replacement (HR 3.42 [95% CI 2.92, 4.02]), with no association observed for hip osteoarthritis. GWAS identified 20 independent loci associated with femorotibial angle, enriched for pathways related to skeletal development, cartilage biology, and endochondral ossification. Post-GWAS analyses demonstrated regulatory effects across fetal and adult joint tissues, supporting life course influences on alignment. Genetic correlation analyses showed shared architecture between femorotibial angle and knee osteoarthritis. Causal analyses suggested that genetic liability to osteoarthritis reduces femorotibial angle (β −0.11 [−0.16, - 0.06]), while evidence for an overall causal effect of femorotibial angle on osteoarthritis risk was limited (OR 0.93 [0.79, 1.10]).

**Conclusions:** Knee alignment and susceptibility to knee osteoarthritis are partially genetically determined. At the population level, these genetic determinants support a causal effect of osteoarthritis on knee alignment, whereas evidence for a causal effect of alignment on knee osteoarthritis was limited. Furthermore, this study identifies novel genetic loci linking knee alignment with pathways involved in skeletal development and cartilage biology relevant to osteoarthritis.

**Key messages:** *What is already known on this topic?:*

- Lower limb malalignment, particularly varus alignment, is associated with knee osteoarthritis.
- Previous studies have been observational, limiting their ability to determine whether malalignment is a cause or consequence of osteoarthritis.
- Genetic influences on knee alignment have been recognized, but its underlying genetic architecture and relationship with osteoarthritis susceptibility remain poorly understood.

*What this study adds?:*

- This study identifies 20 genetic loci associated with femorotibial angle, which demonstrate enrichment in molecular pathways responsible for bone and cartilage development and maintenance across the life course.
- Instrumental variables analyses using genetic data provides greater evidence that osteoarthritis has a causal influence on knee alignment than alignment has on osteoarthritis risk.

*How might this affect research, practice or policy?:*

- At a population level, knee alignment should be considered a marker of disease severity and progression rather than a causal target for osteoarthritis prevention.
- These findings support the use of alignment-modifying interventions, such as osteotomy and bracing, to slow progression in established osteoarthritis but not as a means of preventing disease onset in the general population.
- Novel genetic pathways associated with varus deformity and osteoarthritis could guide future patient stratification and targeted treatment.

## Introduction

Osteoarthritis is the most common form of arthritis and a leading cause of chronic pain and long-term disability^1^. While osteoarthritis can affect any joint, knee osteoarthritis is the most common, with an estimated 368 million cases worldwide^2^. Knee osteoarthritis substantially reduces patients’ quality of life and imposes considerable direct healthcare costs, estimated at $27 billion annually in the United States, representing a major public health burden^3^.

Lower limb alignment is a key determinant of load distribution across the knee joint and disturbances are thought to contribute to the development of knee osteoarthritis^4 5^. Varus alignment, colloquially known as “bow legged”, shifts load medially, increasing compressive force in the medial compartment. Whereas valgus alignment, known as “knock kneed”, displaces load to the lateral compartment^5 6^. The femorotibial angle is often used to quantify alignment and can be evaluated using a range of imaging modalities, including X-ray, CT, and MRI^7–9^. Most studies use standing long-leg X-rays as these are the clinical standard for weight-bearing measurement, but these investigations involve relatively small samples, and are focused on patients about to undergo surgery, where it is difficult to discern cause from effect^10^. Osteoarthritis itself is known to alter knee alignment during progression, with medial compartment disease associated with varus alignment^5 10 11^. In addition, forces transmitted through the hip joint are also thought to vary according to knee alignment, yet their relationship with hip osteoarthritis has been investigated far less extensively^12^.

Determinants of an individual’s knee alignment are poorly understood. At a population level, it is known that females are more prone to valgus alignment, whereas males are more prone to varus alignment and this is thought to be due to sex differences in pelvic shape^13–15^. A previous sibling study estimated the heritability of femorotibial alignment to be around 50%, suggesting an underlying genetic component^16^. This notion is also supported by previous genetic studies of knee osteoarthritis, which have identified genes, such as *GDF5*, that are known to be important regulators of long bone and joint development^17^. The genetic contribution to knee alignment raises the possibility that genetically determined malalignment contributes to susceptibility to knee osteoarthritis^18^.

The UK Biobank is a large, prospective population-based cohort with dual-energy X-ray absorptiometry (DXA) imaging of the whole-body and genetic data available^19^. This resource enables assessment of skeletal morphology and large-scale genome-wide association studies (GWAS)^20 21^. A previous investigation using this dataset proved its utility for understanding the genetics of limb alignment but did not explore their relationships with knee osteoarthritis^12^. In this study, we aimed to (1) derive femorotibial angle in twice the number of UK Biobank participants and investigate its associations with clinical outcomes related to both knee and hip osteoarthritis. (2) Undertake gene-mapping studies across the life course to identify genetic and biological determinants of knee alignment. (3) Use these new genetic data to explore potential causal relationships between femorotibial angle and knee osteoarthritis.

## METHODS

### FTA determination

Lower limb alignment was derived from whole-body DXA images in UK Biobank (Supplementary Methods), using a machine-learning algorithm based on Random Forest regression voting (BoneFinder, The University of Manchester^22^). A total of 73 outline points were automatically placed and equally spaced around the femur and tibia. The femoral and tibial axes were defined as the longitudinal axes passing through the midpoints calculated between corresponding medial and lateral shaft outline points. Subsequently, a custom Python script calculated the anatomic femorotibial angle between the femoral and tibial axes (Figure 1)^23^. The femorotibial angle was classified into severe varus (<0°), varus (0°–2°), neutral (2°–8°), valgus (8°–10°) and severe valgus (>10°), based on previous literature^24–26^. Model performance was evaluated against manual annotations and performed well (Supplementary Methods and Supplementary Table 1). All participants provided informed consent, including linkage to health records. This project was approved under UK Biobank application 17295.

**Figure 1.**
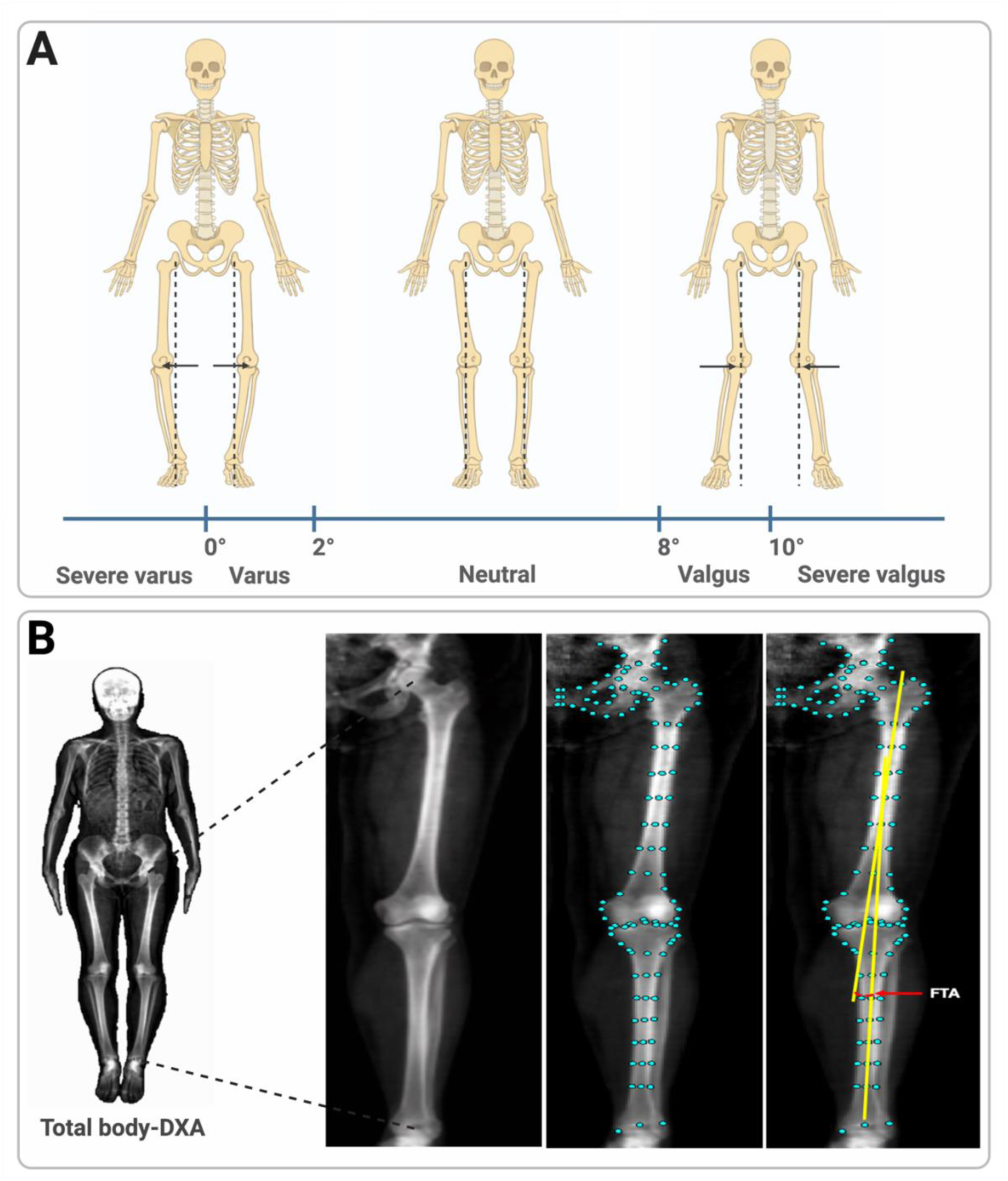
Derivation and classification of the femorotibial angle. (A) Illustration of femorotibial angle classification, ranging from severe varus to severe valgus alignment. (B) Total-body DXA scans were used to derive the femorotibial angle. Automated landmarks were placed along the femur and tibia to define the anatomical axes. The femorotibial angle was calculated as the angle formed between the anatomic femoral and tibial axes (yellow lines). Reproduced by kind permission of UK Biobank ©.

### Observational analyses

Observational associations were calculated between varus and valgus deformities and clinical outcomes related to both knee and hip osteoarthritis. Chronic (>3 months) hip/knee pain was obtained from the UK Biobank touchscreen questionnaire completed on the same day as DXA scan. Hospital-diagnosed knee and hip osteoarthritis were identified through linkage to Hospital Episode Statistics (HES) using ICD-10 codes and total joint replacements were obtained using OPCS-4 codes (Supplementary Table 2). Associations between varus and valgus deformities and clinical outcomes were examined using clustered logistic regression and Cox proportional hazard models (Stata version 18), accounting for within-individual correlation between left and right femorotibial angle measurements. Analyses were conducted at the limb level, with each knee contributing separately. Odds ratios (ORs) or hazard ratios (HRs) with 95% confidence intervals (CIs) were estimated by comparing the odds or hazard of the outcome in exposed knees with those in the control group (Supplementary Methods).

### Genome-wide Association Study of femorotibial angle

Femorotibial angle measurements were standardized to Z-scores (mean=0, SD=1). Subsequently, the mean of left and right measures was calculated and used for GWAS, given that a genetic component of alignment was thought likely to affect both sides simultaneously.

The population was restricted to European ancestry to avoid confounding from differences in genetic ancestry. GWAS was conducted using BOLT-LMM (version 2.3.5) and adjusted for age, sex, genotyping array, and the first 20 genetic ancestry informative principal components^27^. Genome-wide significance was defined as p<5×10⁻⁸. Lead genetic variants were identified using conditional and joint analysis (COJO), with the 1000 Genomes European reference panel^28^.

### Post-GWAS functional and causal analyses

#### Single Nucleotide Polymorphism (SNP) heritability and genetic correlation

SNP heritability of femorotibial angle was estimated using linkage disequilibrium score regression (LDSC v1.0.1), again using the 1000 Genomes European reference panel^29^. LDSC was used to estimate genetic correlations between femorotibial angle and body composition, and musculoskeletal traits (Supplementary Methods).

#### Genetic fine mapping (eQTL, mQTL, ATAC-seq, Bulk RNA-seq)

Functional fine-mapping and effector gene prioritization were performed using multiple post-GWAS analyses: (i) Initially colocalization between 1Mb regions around lead variants for femorotibial angle was conducted with expression quantitative trait loci (eQTL) derived from adult joint tissues, including macroscopically degraded and intact cartilage, synovium, and infrapatellar fat pad obtained from individuals undergoing joint replacement surgery^30^. (ii) Colocalization with methylation quantitative trait loci (mQTL) derived from the same tissue as the eQTL data plus fetal knee cartilage, with samples collected between 7 and 21 post-conception weeks^30 31^. (iii) Chromatin accessibility using ATAC-seq on human fetal skeletal tissues, and human fetal distal femoral cartilage^32 33^, and healthy adult chondrocytes^34^. (iv) Generalized gene-set analysis (MAGMA) also mapped SNP-level association statistics to identify further effector genes before focusing this approach on gene expression programs of bone marrow cells isolated from human femoral head samples^35 36^. (v) Subsequently, the putative effector genes identified by functional genetic analyses (eQTL, mQTL, ATAC-seq) were evaluated across tissues and developmental stages to identify context-specific effects. In addition, these genes were examined using bulk RNA-seq in adult healthy tendon tissue^37^. Pathway enrichment was performed using FUMA (https://fuma.ctglab.nl/) using data from Gene Ontology (GO) and WikiPathways^38 39^. Further details are in the Supplementary Methods.

#### Disease implication

Genes identified through fine mapping were looked-up in three pre-defined gene sets: (i) osteoarthritis-associated genes^40 41^, (ii) developmental genes derived from the limb development atlas^42^, and (iii) skeletal dysplasia genes defined in the nosology of genetic skeletal disorders^43^.

#### Causal analyses

Mendelian Randomization (MR) was used to examine for bidirectional causal effects between femorotibial angle and knee osteoarthritis using the TwoSampleMR package in R^44 45^. Lead variants obtained from COJO were used as genetic instruments, and knee osteoarthritis instruments were obtained from the Genetics of Osteoarthritis consortium^41^. Inverse-variance weighting was the primary analysis, with MR-Egger regression, weighted median and mode-based estimators as sensitivity analyses to account for potential horizontal pleiotropy. To investigate heterogeneous causal mechanisms, we applied MR-Clust to identify clusters of variants with distinct causal effects^46^.

### Patient and public involvement

No patient or public involvement was conducted as part of this study.

## Results

### Population summary

There were 67,613 total-body DXA scans available for analysis at the start of the study, after quality control 66,384 remained (Supplementary Figure 1). The observational analyses were conducted on 54,232 participants with most exclusions (n=9,141) due to the DXA scan being conducted after electronic healthcare linkage ended (Supplementary Methods). The mean age of participants was 64.6 years (range 45–85), with 52.0% female participants. The mean femorotibial angle was 4.8° (range −13.6°–16.8°), with similar values observed for the left and right (4.7° and 4.9°, respectively). The prevalence of neutral alignment was approximately 73%, with the remainder showing varus or valgus alignment. Varus alignment was more prevalent in men, whereas valgus alignment was more common in women, reflecting known sexual dimorphism (Table 1)^13^.

**Table 1.** Descriptive statistics for observational analyses.

|  | Combined | Males | Females |
| --- | --- | --- | --- |
| Participants | 54,232 [100%] | 26,036 [48.0%] | 28,196 [52.0%] |
| **Demographics** | **Mean ± SD [Range]** | **Mean ± SD [Range]** | **Mean ± SD [Range]** |
| Age (years) | 64.6 ± 7.7 [45–85] | 65.3 ± 7.8 [45–85] | 64.0 ± 7.6 [45–84] |
| Weight (kg) | 75.4 ± 15.2 [34–171] | 83.1 ± 13.4 [44–171] | 68.3 ± 13.1 [34–169] |
| Height (cm) | 170.0 ± 9.4 [124–204] | 177.1 ± 6.6 [148–204] | 163.5 ± 6.4 [124–198] |
| **Ethnicity** | **Prevalence [%]** | **Prevalence [%]** | **Prevalence [%]** |
| White | 52,375 [96.6%] | 25,127 [96.5%] | 27,248 [96.6%] |
| Non-white | 1,857 [3.4%] | 909 [3.5%] | 948 [3.4%] |
| **Clinical outcomes** | **Prevalence [%]** | **Prevalence [%]** | **Prevalence [%]** |
| Hip pain (≥ 3 months) | 4,502 [8.3%] | 1,707 [6.6%] | 2,795 [9.9%] |
| Hospital diagnosed hip OA | 926 [1.7%] | 402 [1.5%] | 524 [1.9%] |
| Total hip replacement | 458 [0.8%] | 195 [0.8%] | 263 [0.9%] |
| Knee pain (≥ 3 months) | 8,082 [14.9%] | 3,835 [14.7%] | 4,247 [15.1%] |
| Hospital diagnosed knee OA | 2,234 [4.1%] | 1,187 [4.6%] | 1,047 [3.7%] |
| Total knee replacement | 552 [1.0%] | 263 [1.0%] | 289 [1.0%] |
| **knee alignment angle** | **Mean ± SD [Range]** | **Mean ± SD [Range]** | **Mean ± SD [Range]** |
| Femorotibial angle (mean) | 4.8° ± 2.7° [-13.6° – 16.8°] | 3.9° ± 2.6° [-13.6° – 13.6°] | 5.6° ± 2.4° [-13.4° – 16.8°] |
| Femorotibial angle (left) | 4.7° ± 2.9° [-14.9° – 19.7°] | 3.8° ± 2.9° [-14.9° – 19.7°] | 5.6° ± 2.6° [-13.2° – 17.3°] |
| Femorotibial angle (right) | 4.9° ± 2.9° [-15.9° – 24.2°] | 4.0° ± 2.9° [-15.1° – 24.2°] | 5.7° ± 2.6° [-15.9° – 24.0°] |
| **varus/valgus** | **Prevalence [%]** | **Prevalence [%]** | **Prevalence [%]** |
| Neutral (left) | 39,513 [72.9%] | 18,377 [70.6%] | 21,136 [75.0%] |
| Neutral (right) | 39,860 [73.5%] | 18,616 [71.5%] | 21,244 [75.3%] |
| Varus (left) | 8,548 [15.8%] | 6,251 [24.0%] | 2,297 [8.1%] |
| Varus (right) | 7,744 [14.3%] | 5,699 [21.9%] | 2,045 [7.3%] |
| Severe varus (left) | 3,006 [5.5%] | 2,335 [9.0%] | 671 [2.4%] |
| Severe varus (right) | 2,693 [5.0%] | 2,080 [8.0%] | 613 [2.2%] |
| Valgus (left) | 6,171 [11.4%] | 1,408 [5.4%] | 4,763 [16.9%] |
| Valgus (right) | 6,628 [12.2%] | 1,721 [6.6%] | 4,907 [17.4%] |
| Severe valgus (left) | 1,178 [2.2%] | 205 [0.8%] | 973 [3.5%] |
| Severe valgus (right) | 1,452 [2.7%] | 360 [1.4%] | 1,092 [3.9%] |

Mean refers to the average value of both left and right angles. Neutral indicates knees with alignment values falling within the normal range. SD, standard deviation; OA, osteoarthritis.

### Observational associations

Varus alignment was associated with knee pain, hospital diagnosed knee osteoarthritis, and total knee replacement, with progressively stronger effect estimates (OR 1.73 [95% CI 1.64, 1.82], 2.35 [2.16, 2.56], and HR 3.42 [2.92, 4.02], respectively), while no associations were found with hip outcomes. Stronger associations were seen between severe varus and the same outcomes (Figure 2, Supplementary Table 3). Valgus alignment showed little evidence for an association with either knee pain or osteoarthritis but did show a weak association with total knee replacement. Severe valgus alignment was associated with knee pain, osteoarthritis, and total knee replacement, with effect estimates (1.39 [1.24, 1.56], 1.83 [1.51, 2.20], and 3.09 [2.29, 4.15]) of a similar magnitude to those seen with varus alignment but much weaker as compared with severe varus. Again, there were no associations with hip outcomes (Figure 2, Supplementary Table 3). Sex-stratified associations were broadly consistent with those from combined analyses, with larger effect estimates for varus alignment in women and for valgus alignment in men (Supplementary Figure 2, Supplementary Table 3).

**Figure 2.**
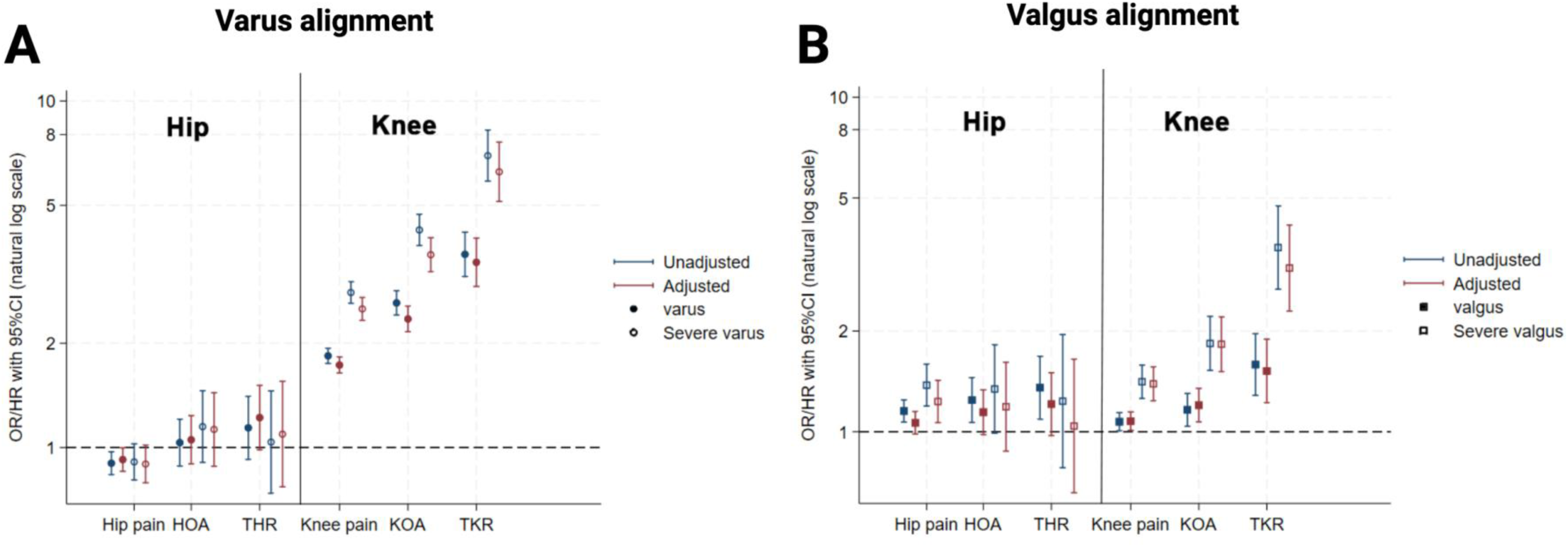
Associations between femorotibial angle and hip and knee clinical outcomes. Odds ratios or hazard ratios with 95% CI are shown for hip and knee pain, hip and knee hospital-diagnosed OA, and total hip and knee replacement, stratified by knee alignment. (A) Varus and severe varus alignment. (B) Valgus and severe valgus alignment. Blue markers indicate unadjusted estimates; red markers indicate adjusted for age, sex, height, weight, and ethnicity (white vs non-white). OR, odds ratios; HR: hazard ratios; CI, confidence Interval; HOA, hospital-diagnosed hip osteoarthritis; KOA, hospital-diagnosed knee osteoarthritis; THR, total hip replacement; TKR, total knee replacement.

### Genome-wide association study

GWAS was performed on the mean femorotibial angle for 58,203 participants of European ancestry (Supplementary Figure 1, Supplementary Table 4). Twenty statistically independent genome-wide significant signals (p< 5×10⁻⁸) were detected (Figure 3A, Supplementary Table 5). The quantile-quantile plot showed evidence of genomic inflation (λ_GC_ = 1.09), however the LDSC intercept term was close to one (*a* = 1.02) suggesting the inflation was mainly due to polygenicity (Supplementary Figure 3).

**Figure 3.**
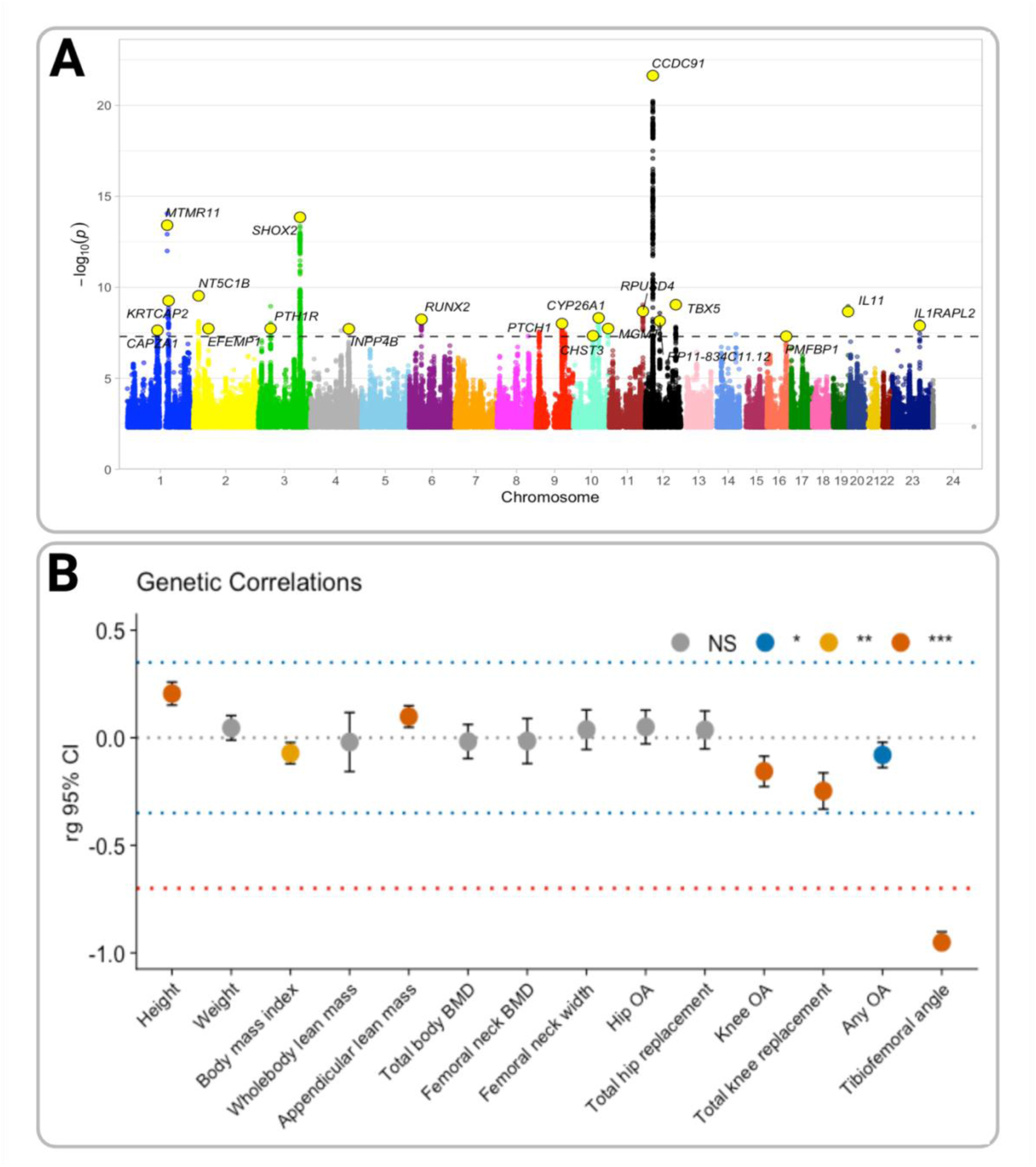
Genetic summary results for femorotibial angle. (A) Manhattan plot showing genome-wide association results for femorotibial angle. Independent lead variants are highlighted in yellow with annotated closest genes. The dotted line indicates the genome-wide significance threshold (p < 5 × 10⁻⁸). (B) Genetic correlations between femorotibial angle and body composition, bone, and osteoarthritis traits. rg estimates are shown with 95% CI, colors denote statical significance levels. CI, confidence Interval; BMD, bone mineral density; OA, osteoarthritis.

### Single Nucleotide Polymorphism (SNP) heritability and genetic correlation

Femorotibial angle was estimated to have a modest SNP heritability (h^2^ = 0.19, [95% CI 0.17, 0.22]). Genetic correlation analyses showed strong evidence of some shared genetic architecture between femorotibial angle and knee osteoarthritis (r_g_= −0.16 [95% CI −0.23, −0.09]) and total knee replacement (−0.25 [−0.33, −0.16]) (Figure 3B, Supplementary Table 6). The negative genetic correlation between femorotibial angle and both knee osteoarthritis and total knee replacement suggests that a varus direction of alignment (i.e., lower femorotibial angle) is related to increased disease risk. Notably, there was little evidence of a genetic correlation with hip osteoarthritis (0.06 [−0.03, 0.15]). A strong genetic correlation was seen between femorotibial angle and tibiofemoral angle (−0.95 [−1.00, −0.90)], the latter is a previously reported but opposite measure of lower limb alignment available in approximately half the current sample size of UK Biobank (n=31,221)^12^.

### Genetic fine mapping

Gene prioritization analyses identified 59 plausible effector genes across the associated loci (Figure 4, Supplementary Table 7). Evidence supporting these genes included eQTL signals in adult joint tissues (n = 2), mQTL in fetal cartilage and/or adult joint tissues (n = 30), overlap with ATAC-seq peaks in fetal knee cartilage (n = 19) and adult talar cartilage (n =14), closest genes to lead variants (n = 20), and genes identified by MAGMA gene-based tests (n = 33). Together, these analyses reflect a broad and integrative approach to candidate gene prioritization. Detailed information on the supporting tissues, techniques and datasets, for each of the prioritized genes are provided (Supplementary Tables 8-9). Once identified, these genes were examined in extra-articular connective tissues (e.g., tendons) using bulk RNA-seq with the top 25% of genes in terms of relative expression highlighted (Supplementary Figure 4, Supplementary Tables 10-11).

**Figure 4.**
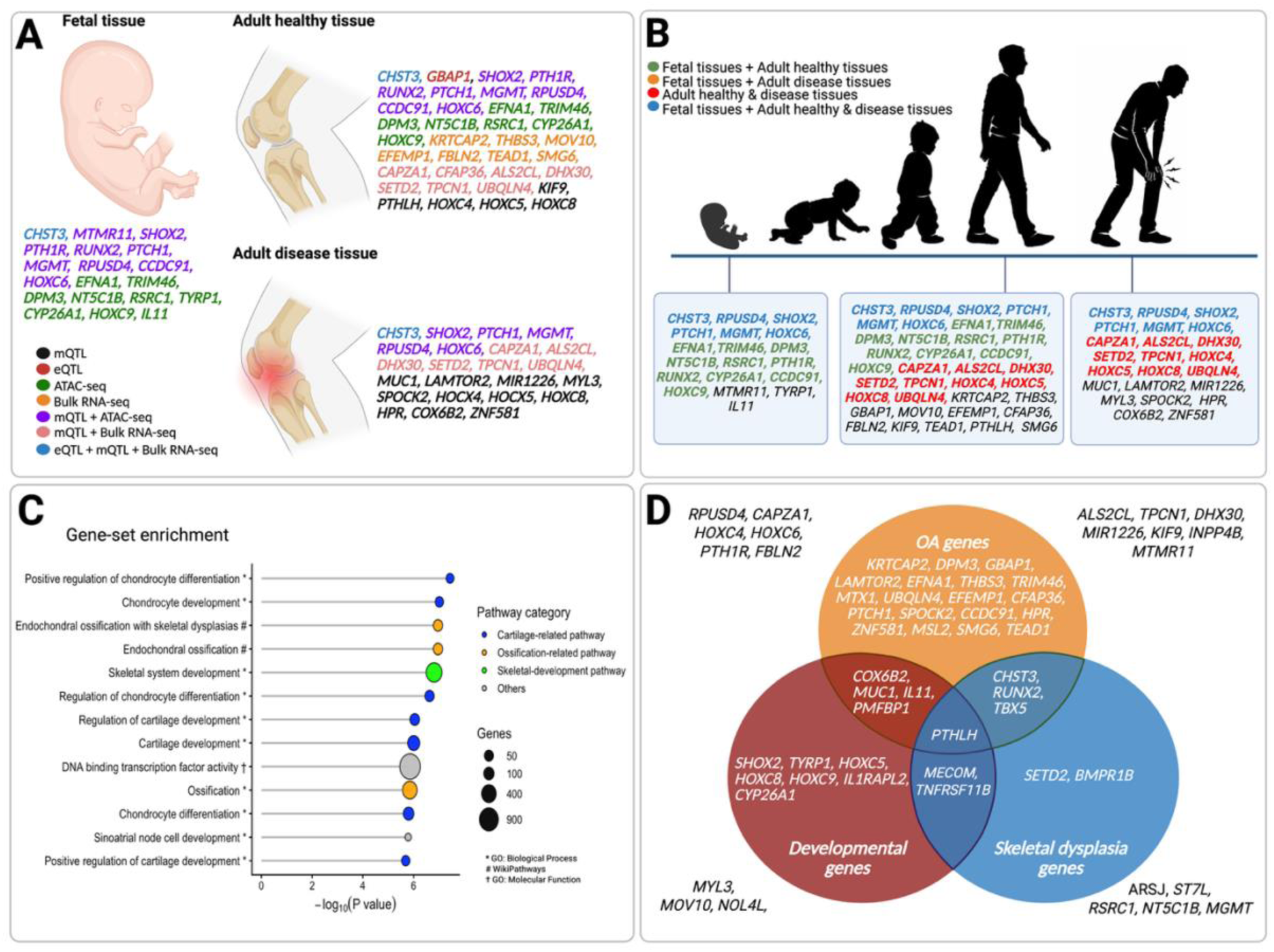
Prioritized genes across tissue contexts, developmental stages, and biological pathways. (A) Prioritized genes across fetal, adult healthy, and adult disease tissue contexts, supported by mQTL, eQTL, ATAC-seq, and bulk RNA-seq evidence. (B) Distribution of prioritized genes across life course. (C) Gene-set enrichment for mapped femorotibial angle-associated genes, illustrating the top enriched biological pathways grouped into cartilage-related (blue), ossification-related (orange), skeletal development (green), and other functional categories (grey). Point size represents gene count; x-axis shows –log₁₀(p-value). (D) Cross-referencing of femorotibial angle-associated genes with previous published osteoarthritis^40 41^, developmental^42^, and skeletal dysplasia^43^ gene sets. eQTL, expression quantitative trait locus; mQTL, methylation quantitative trait locus; ATAC-seq, assay for transposase-accessible chromatin using sequencing.

To further characterize the biological context of these genes, tissue- and stage-specific regulatory annotation and expression analyses were integrated across developmental and adult bone and joint tissues. During this stage we excluded any genes identified through proximity and/or MAGMA alone. The tissue and fine-mapping specific evidence supporting each candidate gene are summarized across tissues (Figure 4A) and the life course (Figure 4B).

### Pathway enrichment

Pathway enrichment revealed biological processes primarily involving chondrocyte differentiation, cartilage development, and ossification-related processes (Figure 4C & Supplementary Table 12). These findings were as expected for a skeletal alignment phenotype and support a developmental and cartilage-bone regulatory context for the prioritized effector genes. Consistent with this, supplementary single-cell RNA sequencing analyses also showed that chondrocyte and fibroblast cells isolated from human femoral heads were enriched for femorotibial angle associated genes (Supplementary Figure 5).

### Skeletal disease mapping

Cross-referencing of the 59 femorotibial angle-associated genes with curated osteoarthritis, developmental limb, and skeletal dysplasia gene sets demonstrated partial overlap across categories (Figure 4D). Twenty-seven femorotibial angle-associated genes, out of the 59, mapped to the osteoarthritis-related gene set. While 21 genes were not represented in the previous studies. Overlap was observed between osteoarthritis and developmental genes, osteoarthritis and skeletal dysplasia genes, and developmental and skeletal dysplasia genes. *PTHLH* was shared across all three categories.

### Mendelian randomization

To reduce the effects of confounding and reverse causation to which the observational studies are often liable, Mendelian randomization analyses to estimate the causal effect of femorotibial angle on knee osteoarthritis risk were performed using the 18 lead variants as genetic proxies (Figure 5A-i). Two variants were excluded as they were not present in the knee osteoarthritis GWAS. Overall, the primary inverse-variance weighting (IVW) analysis showed little evidence of an effect of femorotibial angle on knee osteoarthritis risk (OR per SD increase in femorotibial angle, 0.93 [95% CI 0.79, 1.10]). This was mirrored by sensitivity analyses (Figure 5A-ii, Supplementary Table 13). MR-Clust analysis was applied to account for potential heterogeneity in causal effects across variants with both varus and valgus alignment potentially causing osteoarthritis. This method aims to identify clusters of variants with distinct causal estimates^46^. Among the femorotibial angle associated variants, MR-Clust identified effect, null, and “junk” clusters, indicating heterogeneity in their estimated effects on knee osteoarthritis risk (Figure 5A-iii). A distinct effect cluster was identified, with a negative causal estimate (cluster mean θ: −0.35), suggesting this group of variants leads to a lower femorotibial angle (varus alignment) and causes knee osteoarthritis (Table 2, Supplementary Table 14). These variants included those mapped to *RPUSD4, PTH1R-PTHLH* and *CHST3*.

**Figure 5.**
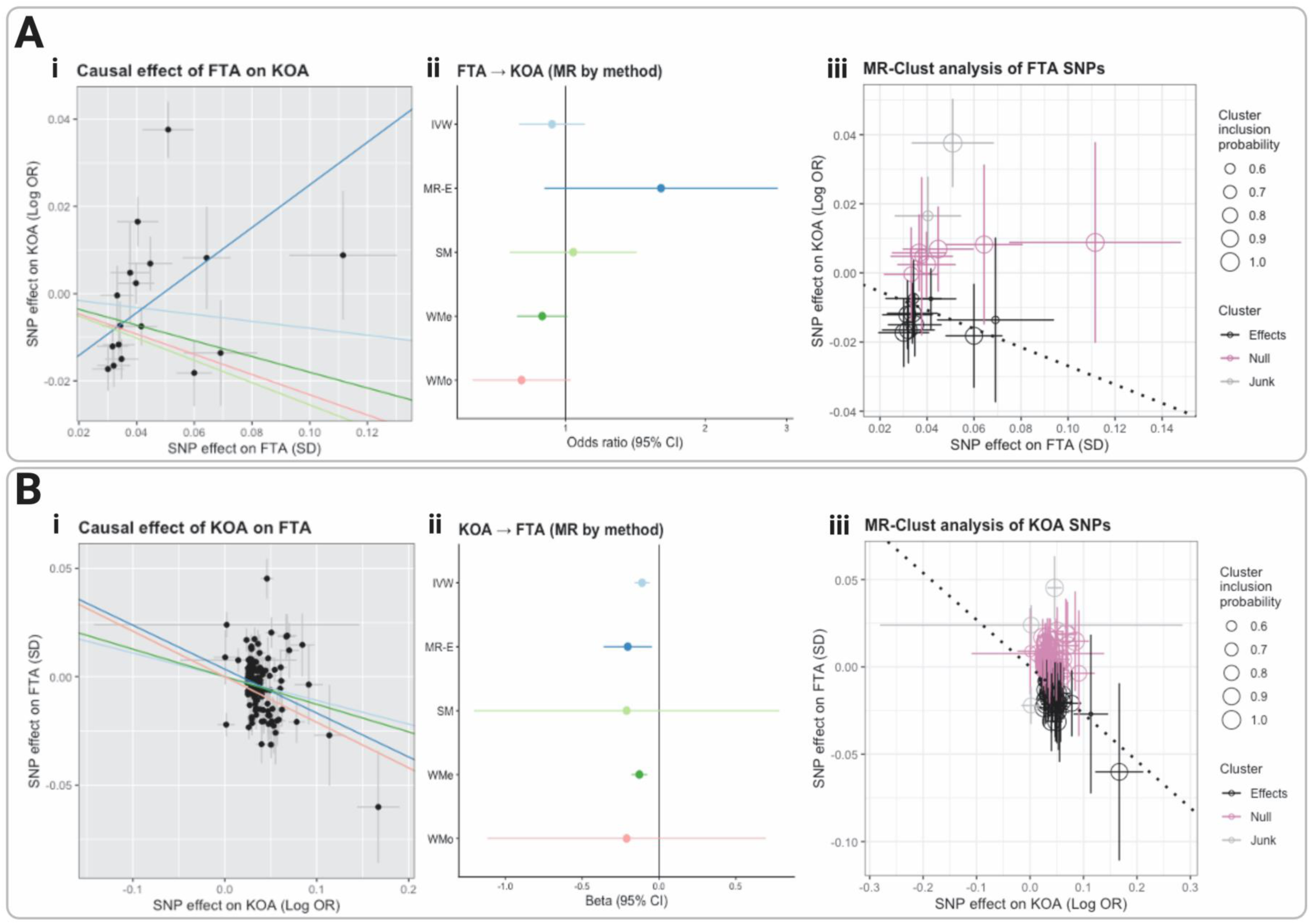
Mendelian randomization (MR) analyses of femorotibial angle and knee osteoarthritis (KOA). (A) MR analyses using 18 independent femorotibial angle-associated lead variants as genetic instruments for femorotibial angle to estimate its causal effect on KOA. (A-i) Scatter plot of SNP effects on femorotibial angle (x-axis) versus knee osteoarthritis (y-axis), with lines representing causal estimates from the different MR methods. (A-ii) Forest plot summarizing causal effect estimates of femorotibial angle on knee osteoarthritis across five MR methods. (A-iii) MR-Clust analysis of femorotibial angle-associated lead variants, showing clustered SNP-specific causal estimates. Point size reflects cluster inclusion probability, and colors denote cluster membership. The dotted line indicates the estimated causal effect for the Effects cluster. (B) Reverse MR analyses using 135 independent knee osteoarthritis-associated lead variants as genetic instruments for knee osteoarthritis to estimate its causal effect on femorotibial angle. (B-i) Scatter plot of SNP effects on KOA (x-axis) versus femorotibial angle (y-axis). (B-ii) Forest plot summarizing causal effect estimates of knee osteoarthritis on femorotibial angle across five MR methods. (B-iii) MR-Clust analysis of knee osteoarthritis -associated lead variants, with point size indicating cluster inclusion probability and colours representing cluster membership. The dotted line denotes the causal effect estimate for the Effects cluster. FTA, femorotibial angle; KOA, knee osteoarthritis; OR, odds ratio; SD, standard deviation; CI, confidence interval; MR, Mendelian randomization; SNP, single nucleotide polymorphism; IVW, inverse-variance weighted; MR-Egger; SM, simple mode; WMe, weighted median; WMo, weighted mode.

**Table 2.** Variants contributing to the clustered effect of femorotibial angle on knee osteoarthritis and their prioritized effector genes.

| RSID | FTA-Beta | FTA-P | KOA-OR | KOA-P | Effector gene |
| --- | --- | --- | --- | --- | --- |
| rs10850306 | -0.03 | $8.99 \times 10^{-10}$ | 1.01 | 0.01 | *TBX5, TPCN1* |
| rs11205303 | -0.04 | $3.84 \times 10^{-14}$ | 1.01 | 0.09 | *MTMR11* |
| rs11606126 | 0.03 | $2.07 \times 10^{-9}$ | 0.99 | $1.18 \times 10^{-3}$ | *RPUSD4* |
| rs1373041 | -0.03 | $1.91 \times 10^{-8}$ | 1.01 | 0.19 | *INPP4B* |
| rs146270697 | -0.07 | $4.89 \times 10^{-8}$ | 1.01 | 0.26 | *PMFBP1, HPR* |
| rs34812000 | 0.06 | $2.31 \times 10^{-22}$ | 0.98 | 0.02 | *CCDC91, PTHLH* |
| rs4683302 | -0.03 | $1.84 \times 10^{-8}$ | 1.02 | $7.24 \times 10^{-4}$ | *PTH1R* |
| rs56368105 | 0.03 | $6.99 \times 10^{-9}$ | 0.99 | 0.02 | *HOXC6, HOXC8, HOXC9* |
| rs6480592 | 0.03 | $4.56 \times 10^{-8}$ | 0.98 | $5.38 \times 10^{-4}$ | *CHST3* |

The variant 12:28407705 reported in the COJO output corresponds to rs34812000 after harmonization and alignment across datasets. FTA, femorotibial angle; OR, odds ratio; P, P-value. KOA-OR and P-value taken from the largest GWAS of knee osteoarthritis<sup>41</sup>.

In the reverse-direction MR, 135 variants, obtained from the Genetics of Osteoarthritis consortium^41^, provided genetic proxies for increased knee osteoarthritis risk (Figure 5B-i). The primary IVW analysis provided strong evidence that genetically predicted knee osteoarthritis has a causal effect on reducing femorotibial angle (β per 1-unit increase in genetically predicted log OR of knee osteoarthritis, −0.11 [95%CI −0.16, −0.06]). Sensitivity analyses showed consistent agreement. These findings suggest that genetic predisposition to knee osteoarthritis causes varus knee alignment. (Figure 5B-ii, Supplementary Table 13). MR-Clust analysis of knee osteoarthritis-associated SNPs similarly identified an effect cluster demonstrating large and consistent causal associations with decreased femorotibial angles (cluster mean θ: −0.34) (Figure 5B-iii, Supplementary Table 15).

## Discussion

This large population-based study provides a comprehensive assessment of lower limb alignment and its relationship with osteoarthritis through the integration of observational, genetic and functional genomic analyses. Strong observational associations were identified between varus alignment and knee osteoarthritis, while genetic analyses provided greater evidence that osteoarthritis influences alignment than alignment influences disease risk. However, a subset of genetic variants linked varus alignment with increased osteoarthritis susceptibility, supporting pathway-specific biological effects rather than a single uniform biomechanical mechanism. Functional genomic analyses identified regulatory signals across fetal and adult joint tissues, implicating pathways involved in chondrocyte differentiation, cartilage biology and endochondral ossification. These findings link variation in limb alignment with biological processes relevant to both skeletal development and osteoarthritis.

Varus alignment demonstrated a substantially stronger association with knee osteoarthritis than valgus alignment, with consistent and progressive associations with knee pain, hospital-diagnosed osteoarthritis, and total knee replacement. This pattern is consistent with the concept that varus alignment is a key biomechanical determinant of medial compartment loading and is more strongly linked to both the presence and severity of disease^5 47^. In contrast, valgus alignment showed weaker associations overall, suggesting that lateral compartment loading plays a less prominent role in population-level knee osteoarthritis risk^48^. The observed sex differences in alignment, with varus more common in males and valgus in females, are consistent with known differences in skeletal morphology, such as wider pelves seen in women^14 15 49^. However, knee alignment likely reflects a combination of anatomical, developmental, and mechanical factors rather than a single determinant^50–52^. Notably, no association was observed between varus or valgus alignment and hip osteoarthritis, suggesting the effects of knee malalignment are largely joint-specific and do not translate to increased risk at the hip. This fits with recent studies that suggest knee and hip osteoarthritis have considerable joint-specific components^53 54^. Although these findings reinforce the strong epidemiological link between malalignment and knee osteoarthritis, the direction of causality remains uncertain. Malalignment may contribute to disease onset through altered load distribution, but it may also arise as a consequence of structural changes such as cartilage loss, meniscal damage and bone remodelling^11^. The stronger associations observed with more advanced outcomes, such as total knee replacement, potentially support a role for malalignment in disease progression^4^.

To build on these observational findings, genetic analyses were applied to provide insight into the association between knee alignment and osteoarthritis, suggesting that this relationship is underpinned by multiple biological mechanisms rather than a single biomechanical pathway^16^. First, the identification of loci enriched for skeletal development and cartilage biology (e.g., *SHOX2*, *TBX5* and *CCDC91*^55–57^) supports a developmental contribution to limb alignment and suggests that variation in skeletal patterning pathways may also influence osteoarthritis susceptibility^31^. Second, Mendelian randomization demonstrated strong evidence that a genetic predisposition to knee osteoarthritis causes a shift towards varus alignment, supporting what is seen clinically, where medial joint space loss and medial compartment overloading during disease progression contribute to varus deformity^11^. Third, there was no evidence that femorotibial angle acts as a uniform causal driver of osteoarthritis risk at the population level. Together, these findings support a model in which much of the variation in knee alignment observed at the population level reflects normal anatomical variation or disease-related remodelling. However, though varus alignment appears to have a limited causal role in the development of knee osteoarthritis in general, a subset of genetic variants link varus alignment with increased disease risk, suggesting that specific developmental and osteogenic pathways may underlie osteoarthritis susceptibility in a subset of individuals^58^.

The integration of functional genomic data provides further insight into the biological mechanisms underlying variation in knee alignment. This is particularly evident for the subset of genetic variants that demonstrate concordant effects on both varus alignment and osteoarthritis risk. The prioritized loci showed evidence across multiple regulatory omics data, including eQTL, mQTL and chromatin accessibility (ATAC-seq) signals in adult cartilage, alongside complementary signals in fetal knee tissues. Signals identified in fetal knee cartilage support a role for early developmental processes in establishing limb geometry, particularly through effects on fetal patterning of the limb (e.g., *HOXC* genes, which are master regulators of morphogenesis^59^) and endochondral ossification (e.g., *MTMR11*, is involved in bone morphogenetic proteins signaling and associated with short stature^60^). In contrast, persistent activity in adult joint tissues suggests that altered expression of these pathways may continue to influence cartilage maintenance and structural integrity during aging and disease. This is supported by genes such as *RPUSD4*, which has been shown to be downregulated in osteoarthritis patients^61^.

Pathway enrichment analyses highlighted processes related to chondrocyte differentiation and cartilage development, which are central to both skeletal patterning and joint homeostasis. Several prioritized genes provide functional support for this link. For example, *CHST3* catalyzes chondroitin sulfation, a process essential for extracellular matrix structure and cartilage integrity^31^. Known mutations in *CHST3* cause monogenic skeletal dysplasia^62^. In this study, *CHST3* showed regulatory evidence across eQTL and mQTL analyses in both adult and fetal joint tissues, and increased expression in tendon tissue. Similarly, *PTHLH* and its receptor *PTHR1* play key roles in chondrocyte proliferation and endochondral ossification and were implicated in femorotibial angle development via ATAC-seq in fetal and adult tissue^63^. These findings suggest that variation in these processes could lead to differences in joint alignment that persist in adulthood. The overlap with these genes and those implicated in osteoarthritis, skeletal development and skeletal dysplasia, particularly *PTHLH*, further supports a shared biological framework linking limb alignment with joint degeneration^42 43 64^.

These findings have important clinical implications for the interpretation and management of knee malalignment. The strong and progressive associations observed between varus alignment and osteoarthritis outcomes reinforce the value of alignment as a marker of disease presence and severity in clinical practice. However, the limited evidence for a general causal effect of femorotibial angle on osteoarthritis risk suggests that malalignment should not be considered a primary universal driver in the initial development of knee osteoarthritis. Instead, the observed reverse relationship indicates that altered alignment seen in osteoarthritis patients often reflects structural changes occurring during disease progression. The extent to which varus malalignment arising during established knee osteoarthritis contributes to subsequent disease progression cannot be determined from our findings. Hence, findings that orthotic and surgical alignment-modifying interventions in patients with established knee osteoarthritis and co-existent malalignment can delay disease progression and improve symptoms are not inconsistent with our results^65 66^. The identification of pathway-specific genetic effects linking alignment and osteoarthritis further raises the possibility that a subset of individuals may be particularly susceptible to alignment-driven disease mechanisms. In this context, improved phenotypic and genetic stratification may help identify patients most likely to benefit from earlier targeted biomechanical and medical interventions.

Several limitations to this study should also be considered. Limb alignment was derived from supine total-body DXA scans, which are more susceptible to positional variation than weight-bearing long-leg radiographs, the current clinical standard^67^. However, the consistency of the observed associations with prior studies using radiographic measures provides reassurance regarding the validity of the phenotype^5 48^. Femorotibial angle was calculated using the anatomical rather than mechanical axis, reflecting the constraints of DXA imaging. In addition, the cross-sectional nature of the imaging data limits the ability to directly assess temporal changes in alignment. Genetic analyses were restricted to individuals of European ancestry, limiting generalisability to other populations; replication in diverse cohorts will be important. Finally, although Mendelian randomization reduces confounding, it remains subject to assumptions including that the genetic proxy affects the outcome only through the exposure and this could be violated by horizontal pleiotropy^44^. That said, the alignment of estimates across sensitivity analyses suggests this was not the case.

In conclusion, knee alignment is a biologically regulated trait that reflects both osteoarthritic remodelling and underlying developmental susceptibility. While strong observational associations support its relevance to disease severity and progression, genetic analyses provide greater evidence that osteoarthritis influences alignment than alignment influences disease risk. These findings indicate that malalignment is unlikely to represent a universal primary cause of knee osteoarthritis but rather often reflects structural changes occurring during disease progression. Functional genomic analyses further implicate developmental and cartilage regulatory pathways linking skeletal patterning with later joint degeneration. Overall, these findings support a model in which knee alignment is largely a consequence of osteoarthritis progression, while also remaining a clinically relevant target for interventions aimed at slowing disease progression. They also highlight biologically distinct pathways through which alignment may contribute to osteoarthritis susceptibility in a subset of individuals.

## Data Availability

The data used in this study were obtained from UK Biobank under Application Number 17295. UK Biobank data are available to bona fide researchers upon application and approval through the UK Biobank Access Management System (https://www.ukbiobank.ac.uk). The data analysed in the current study are subject to UK Biobank access procedures and cannot be shared directly by the authors.

## Acknowledgements and affiliations

This research has been conducted using the UK Biobank Resource under Application Number 17295. We would like to thank all UK Biobank participants. This work was carried out using the computational facilities of the Advanced Computing Research Centre, University of Bristol - http://www.bristol.ac.uk/acrc/. Figures were created in https://BioRender.com.

## Data statement

All UK Biobank data (project number 17295) will be available via their data showcase shortly after publication.

## Author contributions

Authors were involved in the following elements of the study:

- Conceptualization: MJ, FA, TC, JHT, BGF
- Data curation: MJ, FA, CC, SC, SEO, JM, RE, FRS, BGF
- Formal analysis: MJ, FA, CC, SC, SEO, JM, KAF, RE, NM, FRS, JG, NCH, CH, GDS, EZ, SS, TC, SJR, SR, JPK, JHT, TC, CL BGF
- Funding acquisition: BGF, JHT, SS, EZ, CC, JG, RA, NCH, GDS, CH, SS, TC, SJR, TDC, BGF
- Investigation: BGF, MJ, RE, FRS
- Methodology: BGF, RE, FRS, JG, NH, JPK, YZ, GDS, RA, CL, TC, JHT
- Software: FA, RE, CL, TC
- Supervision: BGF, SS, JM, SJR, CC, TC, TDC, CL, SA, CH, NCH, JPK, GDS, RA, JHT
- Validation: BGF, MJ, RE, FRS
- Visualization: BGF, MJ, RE, FRS, KAF
- Writing – original draft: MJ, JHT, BGF
- Writing – review and editing: MJ, FA, CC, SC, SEO, SO, KAF, NM, JM, RE, FRS, JG, RA, SA, NCH, CH, GDS, EZ, SS, TDC, SJR, SR, JPK, JHT, TC, CL, BGF

## Supplementary methods

### Population

UK Biobank is a prospective cohort that recruited 503,317 individuals aged 40–69 between 2006-10. It collected extensive genetic, questionnaire, clinical assessment and linked electronic health record data on each participant. Since 2014, a subset of participants has undergone high-resolution dual-energy X-ray absorptiometry (DXA) imaging of both knees and hips using a GE-Lunar iDXA scanner (Madison, WI) [1]. UK Biobank received ethical approval from the North-West Multi-Centre Research Ethics Committee (11/NW/0382). All participants provided informed consent, including linkage to health records. This project was approved under UK Biobank application 17295.

### Manual validation of automated femorotibial

Model performance was evaluated against manual annotations in 100 randomly selected scans using concordance correlation coefficients (CCC), demonstrating excellent agreements between manual and automated annotations (CCC 0.95; 95% CI 0.92–0.96). Reliability was assessed in the same 100 scans using intraclass correlation coefficients based on the three repeated measurements, demonstrating excellent repeatability for both manual and automated measurements (Supplementary Table 1).

### Observational analyses

The control group for the observational analyses consisted of participants without either varus or valgus deformities. That is the control group had neutral alignment.

Time to event analyses were restricted to participants with DXA imaging acquired before the study end date [30^th^ October 2022], which was the last date of electronic healthcare linkage. They were censored on death. Analysis was adjusted ethnicity categorized as White versus non-White due to the low numbers of non-White participants.

### GWAS

Genotyping, imputation and quality control (QC) were performed by UKB as previously described [2]. Samples were genotyped using two genotyping arrays; Applied Biosystems UK BiLEVE Axiom Array by Affymetrix (49,950 participants) and Applied Biosystems UK Biobank Axiom Array (438,427 participants). Data were imputed using the HRC reference panel, and the merged UK10K and 1000 Genomes phase 3 reference panels in IMPUTE4.

Ancestry assignment of UKB participants was performed as follows: the UKB sample was projected onto the first 20 principal components estimated from the 1000 Genomes Phase 3 (1000G) project (where ancestry was known) using GCTA version 1.93.2. Projections used a curated set of 38,512 LD-pruned HapMap 3 Release 3 (HM3) bi-allelic SNPs that were shared between the 1000G and UKB genotyped datasets (i.e. MAF > 1%, minor allele count > 5, genotyping call rate > 95%, Hardy-Weinberg P > 1×10^−6^, and regions of extensive LD removed). Uniform Manifold Approximation and Projection for Dimension Reduction (UMAP) was used in conjunction with the first 20 principal components to cluster 486,445 individuals using the following parameters: min_dist=0.0001, n_components=3, n_neighbors=45, random_state=10293082. UKB participants that clustered together with the 1000G European sub-populations were manually identified by visual inspection (N=461,920) and used for downstream genetic analyses.

### Genetic correlation - traits

Osteoarthritis phenotypes including hip osteoarthritis, total hip replacement, knee osteoarthritis, and total knee replacement, were obtained from the largest osteoarthritis GWAS [3]. Height and body mass index were obtained from GIANT consortium [4, 5]. Weight, whole-body lean mass, appendicular lean mass, femoral neck width, and tibio-femoral angle were derived from genome-wide association studies conducted in the UK Biobank [6–9]. Femoral neck bone mineral density (FN-BMD) and total-body DXA-derived BMD were obtained from previous research [10, 11].

### Functional genetic data

#### Colocalization

Expression quantitative trait loci (eQTL) colocalization analyses were performed between the associated loci and eQTL signal to identify regulatory variants influencing gene expression. eQTL data were derived from adult joint tissues, including macroscopically highly degraded and low-degraded cartilage, synovium, and infrapatellar fat pad obtained from individuals undergoing joint replacement surgery [12]. Methylation quantitative trait loci (mQTL) colocalization analyses were also conducted to evaluate genetic effects on DNA methylation at CpG sites. mQTL data were obtained from the same adult joint tissues as the eQTL analyses, as well as from fetal knee cartilage samples collected between 7 and 21 post-conception weeks [12, 13]. Evidence of colocalization was considered suggestive where the posterior probability of a shared causal variant (PP4) was ≥0.60 and confirmed where the posterior probability of a shared causal variant (PP4) was ≥0.80. Closest genes were annotated using CpG-to-gene mapping based on Illumina HumanMethylation450K array annotation (hg19), with rsID positions aligned to GRCh37. Where no gene annotation was available, the nearest gene was identified manually based on base-pair position on the chromosome using dbSNP [14].

#### ATAC-seq

Chromatin accessibility was assessed using assay for transposase-accessible chromatin sequencing (ATAC-seq) in fetal skeletal tissues (67 days post-fertilization), fetal knee cartilage (12 post-conception weeks), and adult talar cartilage samples. The closest genes were annotated based on chromosomal base-pair position using dbSNP [14–17].

#### Generalized gene-set analysis

To explore potential links between femorotibial angle-associated genes and bone cell function, we performed MAGMA competitive gene set analyses on single-cell transcriptomics data generated from human femoral heads, which primarily consist of trabecular bone. Firstly, gene-based association tests were conducted using MAGMA software (v1.10) [18] applied to the European femorotibial angle GWAS summary statistics. Patterns of linkage disequilibrium were modelled with a reference sample of ∼50,000 unrelated individuals of European ancestry from the UK-Biobank Study[19]. Genetic variants were annotated to protein-coding genes based on Ensembl gene version GRCh37. A variant was assigned to a gene if it was located within 2 kb upstream of the transcription start site or within 1 kb downstream of the transcription end site. A multi-model approach that combined results from two complementary gene-level association models, was applied: the ‘SNP-wise mean’ model that calculates the mean of χ2 statistics for all genetic variants annotated to a gene, as well as the ‘SNP-wise top’ model that calculates the χ2 statistic for the lead genetic variant annotated to a gene. The aggregated p-value from this approach (p_Multi_) reflects the overall strength of evidence for association between each protein coding gene and femorotibial angle. The threshold to declare statistical significance was determined using *Bonferroni* correction as follows: error rate of one test / number of genes tested = 0.05 / 20 = p< 2.5×10^−3^.

Human single cell RNA sequencing data was obtained from previously published work[19]. In brief, femoral heads were collected from patients undergoing joint replacement surgery. Bone cores were excised and digested. Viable human cells (DAPI^−^) negative for erythroid marker CD235a were sorted into CD45+ (haematopoietic) and CD45^−^ (non-haematopoietic) populations. Cells were processed using the 10x Chromium platform and Chromium Single Cell 3’ v2 Reagent Kit (target 10,000 cells/channel), followed by sequencing on the Illumina NovaSeq 6000 Sequencing System. Raw data were processed with CellRanger (v2.7) and analysed using the Seurat package (v4 and v5)[20]. Log normalisation was applied and low-quality cells filtered (fewer than 300 genes or Mitochondrial Unique Molecular Identifier (UMI) fraction>10%). Dimensionality reduction was performed using 20 principal components derived from the top 3,000 variable genes, and batch correction was applied using Harmony[21]. Clustering was performed using the Louvain Method on shared nearest neighbour graphs and visualised by Uniform Manifold Approximation and Projection (UMAP). Cell types were annotated using canonical markers and cluster-specific gene programs as described in Chai et al[19]. Differentially expressed genes (gene programs) were identified using Seurat FindAllMarkers, requiring log2FC>0.5 and Bonferroni-adjusted p<0.05.

Evidence of enrichment was inferred by testing whether genes comprising each cell-type gene program were more strongly associated with femorotibial angle on average than protein-coding genes not included in the program. Gene set analysis (GSA) used data from gene-based tests of association and accounted for several confounding factors including gene size, gene density, and the inverse of the mean minor allele count in the gene, as well as the log value of these three factors. Post-hoc permutation analysis was conducted on enriched gene-programs using R-scripts supplied with the MAGMA software using default settings[22].

#### RNA-seq

Gene expression was evaluated using bulk RNA sequencing (RNA-seq) data from adult musculoskeletal tissues, including multiple tendon-related tissues (hamstring, posterior tibialis, extensor hallucis longus, Achilles, patellar, quadriceps, supraspinatus, and long head of biceps tendons). Expression levels were quantified as log_2_counts per million (log_2_CPM), and genes were ranked based on mean expression across tissues. The top 25% of genes were defined as relatively highly expressed, with the top 10% representing a more stringent high-confidence subset, based on relative expression levels within this dataset rather than absolute expression across tissues [23].

